# Long-term clinical and patient reported outcomes of an enhanced monofocal intraocular lens

**DOI:** 10.1101/2024.10.12.24315376

**Authors:** Catharina Latz, Annika Licht, Katharina A. Ponto, Johannes Menzel-Severing, David P. Piñero, Alireza Mirshahi

## Abstract

**Purpose:** To evaluate long-term clinical and patient-reported outcomes (PROMs) following the implantation of an enhanced monofocal intraocular lens (IOL).

**Methods:** This ambispective non-comparative single-centre study involved 41 patients (ages 48-84) who underwent bilateral cataract surgery with the Tecnis Eyhance IOL (model ICB00, Johnson & Johnson Vision). Distance and intermediate visual acuities, refraction, and PROMs were assessed 18 months or more after surgery. Spectacle independence was evaluated using the PRSIQ questionnaire, with patients self-reporting on visual quality, difficulties in performing specific tasks and perception of photic phenomena at distance and intermediate vision.

**Results:** At 18 months or later, 100.0%, 73.2%, 100% and 79.5% of patients achieved a binocular uncorrected distance, uncorrected intermediate, corrected distance, and distance-corrected intermediate visual acuity of 0.20 logMAR or better, respectively. Less than 10% of patients reported photic phenomena. Mean visual quality scores were 1.68±0.72 for distance and 2.05±0.92 for intermediate vision (1=very good to 6=very poor). The dashboard was clearly visible while driving for 95.1% of patients, while 45.0% could perform screen work without glasses; an additional 40.0% could do so with enlarged fonts. Complete spectacle independence was reported by 87.8% for distance vision, and 53.7% for intermediate vision. At least moderately satisfied were 90.2% with distance vision, 87.8% with intermediate vision, and 51.2% with near vision.

**Conclusions:** The enhanced monofocal IOL ICB00 provides good long-term distance and intermediate visual quality, leading to considerable spectacle independence and patient satisfaction. Most patients required near vision correction.

## Introduction

Presbyopia-correcting IOLs can be categorized by their underlying optical-physical properties into diffractive and non-diffractive IOLs. While diffractive IOLs split incoming light through surface discontinuities, non-diffractive IOLs bend light by surface curvature changes. Another way to categorize IOLs is the range of focus they offer. Full range of focus (FROF) IOLS cover vision from distance to near, including intermediate distances. Due to their typically diffractive technology, PROF IOLs may compromise visual experience through dysphotopsia. Partial range of focus (PROF) IOLs cover a partial spectrum of vision, typically extending from distance to intermediate, with some capacity for near vision. They may be diffractive and non-diffractive and create fewer visual compromises such as dysphotopsias. Most recently, Fernandez et al. offered a systematic approach to categorize IOL types based on the shape of the defocus curve: PROF IOLs were divided into narrow, enhanced and extended PROF IOLs^1^.

In 2018, the American National Standard Institute (ANSI) provided specific criteria to define enhanced depth of focus (EDOF) IOLs as IOLs that correct aphakia, with an extended range of focus above a defined functional visual acuity threshold of 0.2 LogMAR, providing useful distance and intermediate vision with monotonically decreasing visual acuity from the best distance focal point.^2^ With the introduction of a new technology named Tecnis Eyhance (model ICB00, Johnson & Johnson Vision) in 2019, a new category of intraocular lenses (IOLs) was generated, popularly known as mono-EDOF, or enhanced monofocal IOLs.^3^ According to Fernandez et al^1^ this IOL would be classified as enhanced PROF. This IOL extends the depth of focus similarly to EDOF IOLs, but does not meet the four effectiveness end-points required to classify it as true EDOF lenses according to the criteria of the American National Standard Z80.35– 2018 (ANSI).^2^ Enhanced monofocal IOLs include some optical modifications to provide an efficacious correction for distance vision while providing an enhanced intermediate visual function. Various studies have been conducted to characterize the clinical performance and patient acceptance of the Eyhance IOL and other enhanced monofocal IOLs.^3–5^ Notably, no higher incidence of postoperative photic phenomena was reported with these IOLs compared to regular monofocal lenses.^6^

The Eyhance ICB00 IOL has an aspherical posterior surface and a modified aspheric anterior surface with a continuous increase in power from the periphery to the center of the lens while maintaining the distance image quality.^7^ The overall lens design is refractive and leverages the geometry, material, and corneal spherical aberration correction features, with a higher order aspheric profile included on the anterior optic surface. As this lens is designed to retain the benefits of a monofocal IOL while adding intermediate vision, the ICB00 IOL is considered an “enhanced” monofocal IOL designed to provide improved intermediate vision.^7^ A variety of clinical studies have demonstrated that this IOL provides good distance visual acuity with enhanced intermediate visual acuity.^8–27^ Specifically, enhanced monofocal IOLs provide comparable results in terms of distance vision compared to conventional monofocal IOLs.^8–27^ However, the number of studies about patient-reported outcomes (PROMs), which reflect the real perception of the patient regarding the improvement achieved with the implantation of the IOL, is still limited.^7,11,17,27^ Furthermore, long-term real-world data is still scarce. Given that neuroadaptation and habituation are the basis for patient satisfaction with all refractive IOLs and that this process takes 6 months and longer, it is of particular interest to analyze patient-reported outcomes at long term. Also, to satisfactorily manage patient expectations, it is especially relevant to know the real level of spectacle independence achieved with this type of enhanced monofocal IOL. The aim of the current study was to evaluate the long-term clinical and patient-reported outcomes after the implantation of the enhanced monofocal IOL ICB00 with special emphasis on the achieved level of spectacle independence. In addition, patient satisfaction with this type of IOL in a real-world setting was analyzed.

## Methods

### Patients

This was an ambispective, non-comparative, single-centre study enrolling a total of 41 patients who underwent uncomplicated phacoemulsification bilateral cataract surgery with implantation of the enhanced monofocal IOL ICB00 and had a follow up visit of 18 months or longer. Recruitment stared on October 17,2023 and ended November 30^th^ 2023. The last patient was examined on December 12^th^, 2023. Included were patients with both eyes having a corneal astigmatism below 0.75D, aged 45 or older, and visually significant cataract. Exclusion criteria included known systemic diseases with the potential of altering the outcome of the study, previous ocular surgery including refractive surgery, irregular astigmatism, zonular alterations that may affect IOL position and stability, active ocular disease, previous diagnosis of retinal pathologies and severe glaucoma, as defined by mean deviation deficits of more than 12 dB on visual fields.

Before inclusion in the study, each patient was informed in detail about the nature of the study, and written informed consent was given according to the tenets of the Declaration of Helsinki. Minors were not included in the study. This study was approved by the medical ethics committee of the Medical Chamber of North-Rhine, Germany (No: 2023012).

### Clinical Protocol

All patients underwent a complete preoperative examination, including measurement of uncorrected distance visual acuity (UDVA) and corrected distance visual acuity (CDVA), objective refraction by autorefractometry, optical biometry and keratometry (IOLMaster 700, Carl Zeiss Meditec), non-contact tonometry, slit lamp biomicroscopy, optical coherence tomography of the macula and optic disc (Carl Zeiss Meditec), and dilated fundus evaluation. In all cases, the Barrett TK formula was used for IOL power calculation, targeting emmetropia.

Postoperatively, patients were evaluated at 1 day and later at the discretion of the referring physician. Eighteen months after cataract surgery, all patients were contacted by phone and/or email and scheduled for a long-term follow-up visit. At this visit, a complete visual evaluation was performed, including the following tests: monocular and binocular measurement of UDVA and CDVA, manifest refraction, and measurement of uncorrected intermediate visual acuity (UIVA) and distance-corrected intermediate visual acuity (DCIVA) (measured at 66 cm). Spectacle independence was evaluated using the Patient-Reported Spectacle Independence Questionnaire (PRSIQ).^28^ We assessed the patient-reported quality of vision and photic phenomena using a self-developed questionnaire, where patients quantified the level of distance and intermediate visual quality on a scale from 1 (excellent) to 6 (extremely poor). Additionally, patients were asked about the perception of halos, glare, blurring, and starbursts at distance and intermediate vision.

### Surgery

All surgeries were performed by two experienced surgeons (AM, KT) using a standard technique of sutureless microincision phacoemulsification. Before surgery, patients received a peribulbar block (4 mL 0.75% bupivacaine, 2 mL 2% mepivacaine and 75 IE hyaluronidase (ESTEVE Pharmaceuticals GmbH) and dilating eye drops (phenylephrine hydrochloride 5%, Ursapharm Arzneimittel GmbH; tropicamide 0.5%, Pharma Stulln GmbH). Patients who were taking warfarin with a high international normalized ratio received topical anesthesia.

A clear corneal incision with a width of 2.4 mm was placed either superior or temporal as well as two paracenteses (1.0 mm). A manual capsulorhexis was performed under ophthalmic viscoelastic device (OVD). Care was taken to achieve a capsulorhexis diameter of approximately 5 mm to ensure complete coverage of the IOL optic with the anterior capsule. Nuclear disassembly and cortical aspiration were performed using the Centurion vision system (Alcon). The IOL was delivered either under irrigation or viscoelastic protection with an injector provided by the manufacturer. At the end of the procedure, 1 mg cefuroxime and 2mg dexamethasone [4 mg/mL] were administered intracamerally and subconjunctivally, respectively.

Postoperative treatment included a combination eye drop four times a day containing dexamethasone, neomycin sulfate, and polymyxin-B-sulfate with ointment at night or, in case of allergies to preservatives, ofloxacin and dexamethasone eyedrops four times a day. This treatment was tapered over 4 weeks.

### Statistical analysis

Data analysis was performed using the software SPSS version 22.0 for Windows (SPSS). Normality of all data distributions was initially evaluated by means of the Kolmogorov-Smirnov test. A descriptive analysis of all continuous variables was carried out, calculating the average values with their corresponding standard deviations and the ranges of maximum and minimum values. For categorical variables, frequencies of different conditions or aspects were determined. A p-value below 0.05 was considered statistically significant.

## Results

### Patient population

A total of 82 eyes of 41 patients with a mean age of 69.4 years (SD: 9.0, median: 70.0, range: 48 to 84 years) was enrolled. The sample comprised 24 males (58.5%) and 17 females (41.5%).

### Refractive outcomes

Table 1 summarizes the visual and refractive outcomes obtained at the last postoperative visit, more than 18 months after surgery: Mean postoperative binocular logMAR UDVA and UIVA values were 0.05 ± 0.07 and 0.18 ± 0.12, respectively.

**Table 1.** Long-term (18 months or longer) postoperative visual and refractive outcomes: CDVA, corrected distance visual acuity; DCIVA, distance-corrected intermediate visual acuity; SD, standard deviation; SE, spherical equivalent; UDVA, uncorrected distance visual acuity; UIVA, uncorrected intermediate visual acuity.

| VA (LogMar) | Right eye | Left eye | Binocular |
| --- | --- | --- | --- |
|  | Mean (SD) | Mean (SD) | Mean (SD) |
|  | Median (Range) | Median (Range) | Median (Range) |
| **UDVA** | 0.11 (0.10) | 0.10 (0.09) | 0.05 (0.07) |
|  | 0.10 (0.00 to 0.30) | 0.10 (0.00 to 0.30) | 0.00 (0.00 to 0.20) |
| **CDVA** | 0.05 (0.08) | 0.04 (0.05) | 0.02 (0.04) |
|  | 0.00 (0.00 to 0.30) | 0.00 (0.00 to 0.20) | 0.00 (0.00 to 0.10) |
| **UIVA** | 0.27 (0.15) | 0.24 (0.14) | 0.18 (0.12) |
|  | 0.30 (0.00 to 0.70) | 0.20 (0.00 to 0.60) | 0.20 (0.00 to 0.50) |
| **DCIVA** | 0.22 (0.13) | 0.22 (0.13) | 0.17 (0.11) |
|  | 0.20 (0.00 to 0.50) | 0.20 (0.00 to 0.50) | 0.20 (0.00 to 0.40) |
| **SE (D)** | 0.06 (0.47) | 0.19 (0.53) | --- |
|  | 0.00 (-0.75 to 1.13) | 0.25 (-1.00 to 1.50) |  |

Likewise, a mean binocular logMAR DCIVA value of 0.17 ± 0.11 was measured. 100.0%, 73.2%, 100% and 79.5% of patients achieved a binocular UDVA, UIVA, CDVA and DCIVA of 0.20 logMAR or better, respectively (Figure 1).

**Figure 1.**
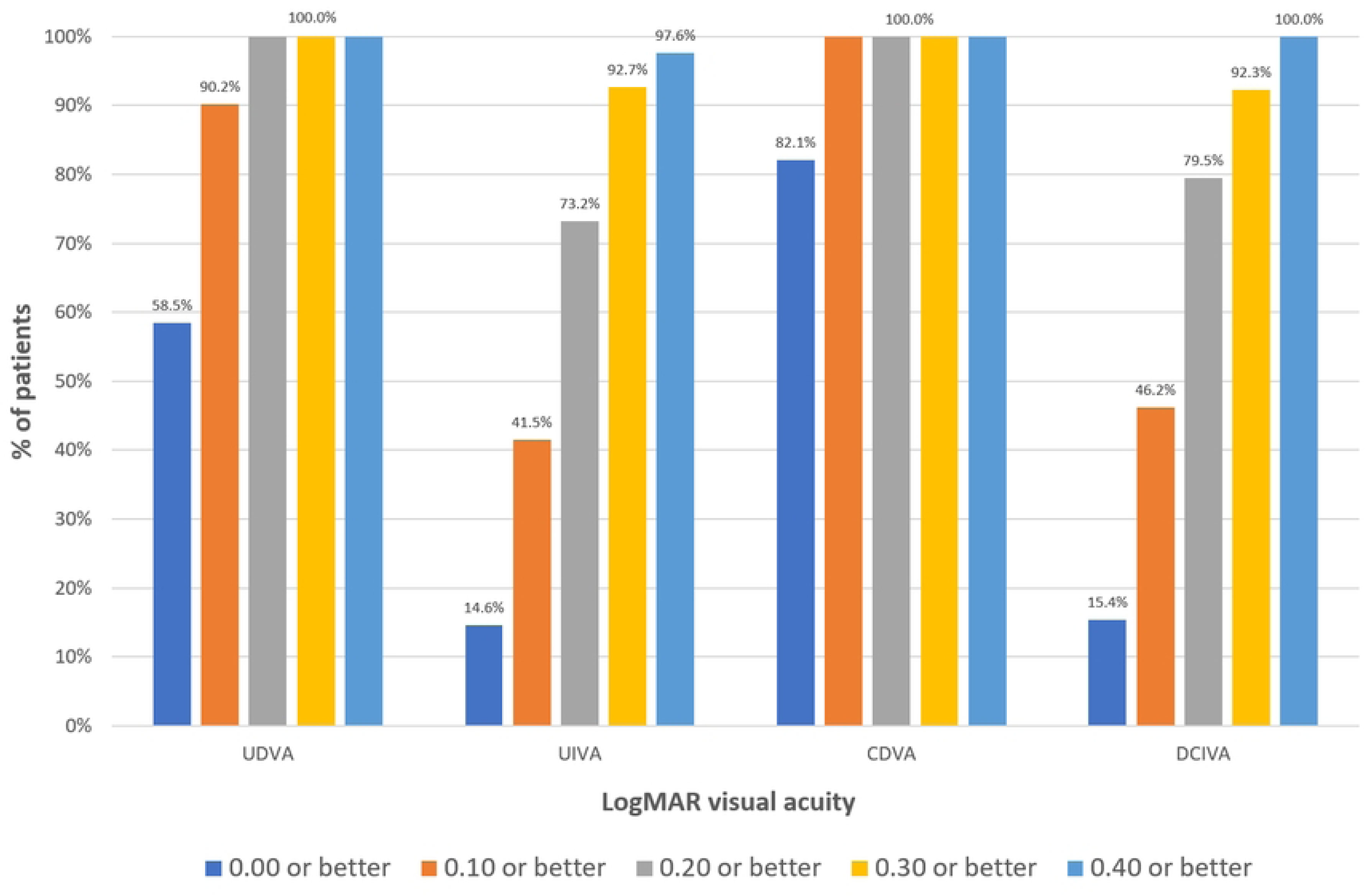
Postoperative binocular visual acuity: uncorrected distance (UDVA), uncorrected intermediate (UIVA), corrected distance (CDVA) and distance-corrected intermediate (DCIVA).

The postoperative spherical equivalent (SE) was within ±0.50 D in 80.5% of right eyes and 82.9% of left eyes. Likewise, postoperative SE was within ±1.00 D in 95.1% and 92.7% of right and left eyes, respectively.

### Patient-reported outcome measurements

PRSIQ and our own developed questionnaire were used to assess PROMs: A small percentage of patients reported postoperative perception of photic phenomena. Specifically, 9.8%, 2.4% and 9.8% of patients reported the perception of halos, blurry vision, and glare at distance vision, respectively (Figure 2).

**Figure 2.**
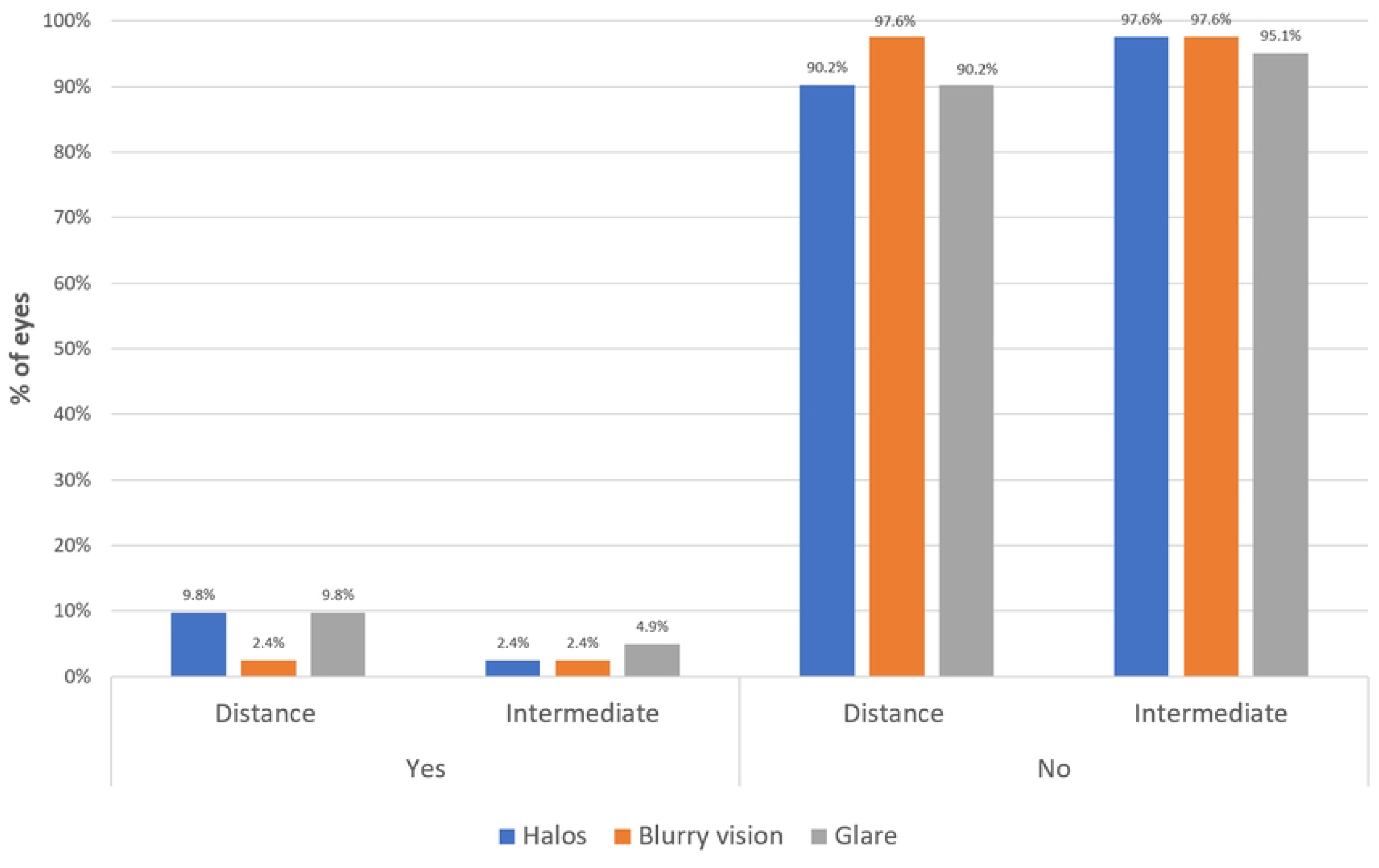
Perception of photic phenomena 18 months postoperatively.

These percentages decreased to 2.4%, 2.4% and 4.9% for the perception of halos, blurry vision, and glare at intermediate vision, respectively (Figure 2). Analysis of this subgroup did not reveal a significantly increased refractive error.

The visual quality achieved at distance and intermediate vision was subjectively graded by the patients on a scale from 1 (very good) to 6 (very poor). This grading scale, adapted from the German school system, is widely accepted and familiar to patients. Mean distance and intermediate visual quality satisfaction scores were 1.68 (SD: 0.72; Median: 2.00; Range: 1 to 3) and 2.05 (SD: 0.92; Median: 2.00; Range: 1 to 4), respectively. No patient provided scores of 4 or worse when asked about satisfaction with distance and intermediate visual quality. A total of 95.1% of patients reported a clearly visible dashboard when driving a car. When the dashboard was brightly illuminated, an additional 97.5 % of patients reported clear visibility. 45% of patients were able to perform screen work without spectacles, 40% had to enlarge the font and 63.4% of patients achieved spectacle-free reading when the font size was large enough.

Regarding the level of spectacle independence in the past 7 days, most patients did not need spectacles for distance vision (87.8%) after surgery, whereas more than half of the sample evaluated did not need them for intermediate vision (53.7%) (Figure 3A).

**Figure 3.**
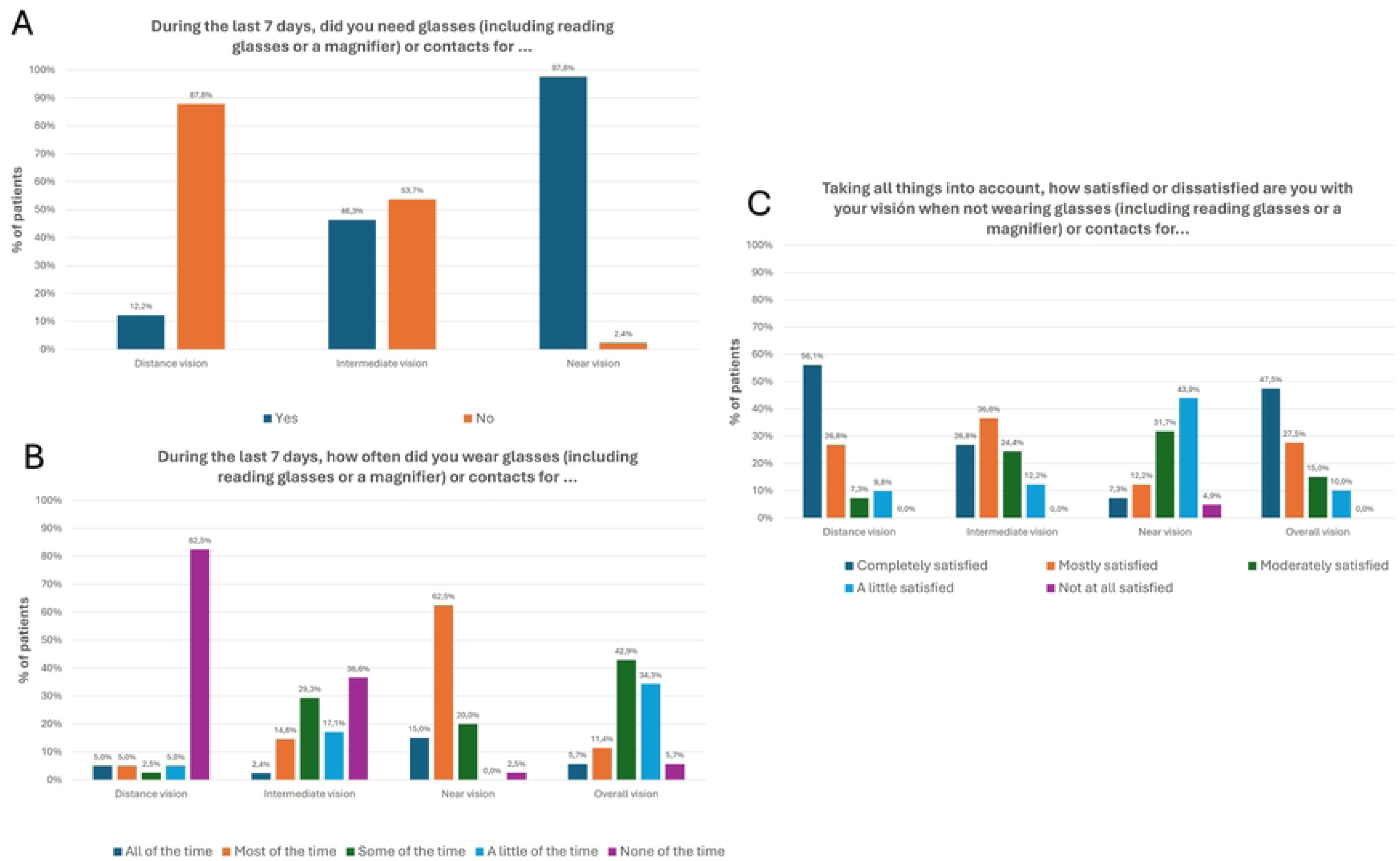
Outcomes obtained with the PRISQ questionnaire.

In contrast, most patients required glasses for near vision activities (97.6%) (Figure 3A). Furthermore, a total of 82.5% of patients did not wear glasses at any time for distance vision during the last 7 days, whereas this percentage decreased to 36.6% for intermediate vision (Figure 3B).

Concerning patient satisfaction, a total of 90.2%, 87.8%, 51.2% and 90.0% of patients were completely, mostly or moderately satisfied with their unaided distance, intermediate, near and overall vision, respectively (Figure 3C).

No adverse events were recorded during the follow-up, with no cases showing a development of posterior capsular opacification (PCO) requiring YAG capsulotomy.

## Discussion

To the best of our knowledge this is the first study to demonstrate excellent distance and good intermediate visual acuities at long term after implantation of the ICB00. No adverse events were noted. Special emphasis was placed on patients’ perception: photic phenomena were denied for distance or intermediate vision by over 90% of study participants. Furthermore, 75% of patients were completely or moderately satisfied with their overall vision. This is consistent with other studies evaluating the same IOL.^7,10,11,17,25,26^ and other models of enhanced monofocal IOLs.^29,30^ Goslings et al^11^ reported mean binocular UDVA of 0.11 ± 0.11, and UIVA of 0.12 ± 0.11 at three months. These visual results are slightly worse than those in our study, which could be explained by a different IOL calculation formula, not specified in the publication. Giglio et al. reported mean postoperative binocular UDVA, UIVA and DCIVA values of -0.03 ± 0.07, 0.17 ± 0.12 and 0.13 ± 0.11, in 30 eyes using the Barrett Universal II formula.^7^ Mencucci et al. also reported similar mean binocular UDVA, UIVA and DCIVA values (0.03 ± 0.05 vs. 0.16 ± 0.10 vs. 0.15 ± 0.08) in 80 eyes of 40 patients using the Holladay 1 formula for axial lengths between 22.0 mm and 25.0 mm and Hoffer Q formula for axial lengths equal to or less than 22.0 mm. Their axial length measurements were obtained using the IOL master 500 (Carl Zeiss Meditec AG)^26^ However, other authors have reported worse UDVA and better UIVA values in eyes implanted with the ICB00 in which a micro-monovision approach had been applied or a trend towards a low myopic residual refractive error was found. ICB00 allows for efficient restoration of distance visual acuity, with enhanced intermediate visual function.

Furthermore, we analyzed how this functional intermediate vision was perceived by patients. Specifically, we investigated the level of spectacle independence, perceived difficulties in daily vision-related tasks, and the perception of photic phenomena. Spectacle independence was evaluated using the PRSIQ questionnaire, a validated patient-reported measure assessing spectacle independence following cataract surgery.^28^ To this date and to the best of our knowledge, our study is the first to report the level of spectacle independence achieved with the model ICB00 at 18 months or later. As is common after cataract surgery with the implantation of any conventional monofocal IOL, the level of spectacle independence at distance was high: 87.8% of patients were spectacle or contact lens free at any time of the day. However, for intermediate vision only 53.7% of patients were spectacle or contact lens free. On the other hand, when examining the degree of spectacle dependence for intermediate vision, only 21.9% of patients used spectacle correction all the time or most of the time. Regarding near vision, the level of spectacle dependence was significant, with 97.6% of patients wearing glasses for such purposes and 75.6% of them wearing them all the time or most of the time.

Using the PRSIQ tool, Stodulka and Pracharova^31^ investigated another IOL with an optical principle comparable to the IOL type we tested, where a special geometry creates a power gradient from the centre to the periphery. Spectacle independence with this EDOF IOL was achieved in over 80% of patients at distance and intermediate vision.

In our study, 90.2% of patients were satisfied with their uncorrected distance and 87.8% with their uncorrected intermediate vision. This confirms the ability of the enhanced monofocal IOL model ICB00 to provide satisfactory distance and intermediate visual outcomes. This is highly relevant since intermediate vision is essential for the use of computers or equivalent handheld electronic devices such as smartphones and tablets.^32^ Regarding near vision, despite the limitations in terms of visual acuity, approximately half of the sample reported being satisfied with their near visual functionality. This can be explained by the visual acuity provided, which might allow a variety of near vision activities without spectacle correction. Indeed, reading activity after surgery was possible in 14.6% of patients without any additional optical aid, and 63.4% of patients were able to read without correction if font size was large enough. This finding is supported by studies by Goslings et al^11^ and Giglio et al^7^, who used the validated questionnaire Catquest 9SF to investigate difficulties in performing different vision-related activities after the implantation of the ICB00 IOL and detected a trend towards improvement in Rasch-calibrated scores of questions about near vision. Lopes et al^17^ evidenced significant differences between eyes implanted with a conventional monofocal IOL and those implanted with the model ICB00 in the level of difficulty in reading newspaper print and reading the prices of goods while shopping, with better outcomes in the group of eyes implanted with the enhanced monofocal IOL. In our sample, screen work could be performed postoperatively without correction by 45.0% of patients, with an additional percentage of 40.0% of patients able to do it without problems if the font was large enough. Likewise, 95.1% of patients could clearly see the dashboard while driving a car. These outcomes are consistent with those obtained with the validated questionnaire Catquest 9SF in other studies evaluating the ICB00, in which the benefit in intermediate vision with the enhanced monofocal IOL over a conventional monofocal was consistently perceived by patients.^7,11,17^

In accordance with all these PROMs, the level of visual quality graded subjectively by the patient for distance and intermediate vision was good or very good for all patients, with no patient reporting poor distance or intermediate visual quality. This was also consistent with a low percentage of patients perceiving photic phenomena, including halos, blurry vision, and glare, at distance and intermediate vision (less than 10% in all cases). This aligns with optical simulations that demonstrated fewer halos with the enhanced monofocal IOL evaluated than with the other two types of extended range of vision IOLs.^33^ Lee and colleagues^34^ compared the enhanced monofocal IOL evaluated in the current study with a diffractive EDOF IOL and found that although spectacle independence was higher in the diffractive group, this was at the expense of more glare and halos. Similarly, Corbelli et al^16^ found that the enhanced monofocal IOL was not inferior to a diffractive IOL regarding intermediate visual outcome and spectacle independence but had the advantage of less perception of halos and glare.

Limitations of our study include the lack of a control group with a monofocal IOL as well as data on corneal spherical aberration of the study patients. Given that neuroadaptation and habituation take 6 months and longer, this study provides important insights into patient-perceived outcomes at long term. In view of a growing array of FROF and PROF IOLs, long term data are very valuable to improve patient satisfaction and patient counselling.

In conclusion, ICB00 provides excellent levels of distance and good intermediate visual quality, leading to satisfactory levels of spectacle independence and patient satisfaction at long term. Increased long-term side-effects or increased photic phenomena were not detected. Near visual outcome, however, was more limited and varied considerably among subjects.

## Data Availability

All relevant data are within the manuscript and its Supporting Information files.

